# Quantifying myocardial repolarization instability using wearable radar-based non-contact estimation of the JT variability index

**DOI:** 10.64898/2025.12.02.25341506

**Authors:** Zainab Riaz, Stephanie I Pella, Neil Tom, Mark Halaki, Tuguy Esgin, Martin Ugander, Mehmet R Yuce

**Author notes:** Contributed equally as first author (Zainab Riaz, Stephanie I Pella). Contributed equally as last author (Martin Ugander, Mehmet R Yuce).

## Abstract

**Background and Aims:** Myocardial repolarization instability measured by electrocardiography (ECG) as the QT variability index (QTVI) predicts arrhythmic risk. We aimed to introduce and validate a non-contact wearable radar-based method for assessing myocardial repolarization instability.

**Methods:** QTVI by ECG (normalized QT variability to normalized heart rate variability) was compared to the analogous J-point-to-T-end variability index (JTVI) by ECG. Simultaneous 2-minute ECG and wearable radar recordings were analysed using an intelligent algorithm to extract mechanical correlates of the J-point and T_end_ from radar-derived cardiac motion signals, and compared to JTVI by ECG.

**Results:** Based on a selection of publicly available datasets of 9 patients with atrial fibrillation, 11 patients with sinus rhythm and 20 healthy people (n=40, 23% with atrial fibrillation, mean±SD heart rate 85±12 beats/min, QTVI −0.36±0.75, JTVI −0.10±0.75), the SD of JT and QT intervals strongly agreed (R²>0.99, bias −0.4±1.5 ms, 1.1±3.9%), and JTVI and QTVI correlated closely (R²=0.98, QTVI=1.0*JTVI−0.26, bias −0.26±0.11). Among a separate group of healthy volunteers (n=20, age 43±11 years, 25% female, heart rate 66±14 beats/min, JTVI - 0.52±0.51), there was excellent agreement for ECG-derived and radar-estimated SD of JT (R²=0.88, bias 0.1±4.9 ms, 6.4±2.7 %), SD of beat-to-beat interval (SDNN) (R²=0.93, bias 16.0±22.1 ms, 9.5±4.9 %), and JTVI (R²=0.99, bias 0.00±0.05, 0.0±6.5 %).

**Conclusion:** QTVI and JTVI adjusted for a small bias are effectively interchangeable for quantifying myocardial repolarization instability, and JTVI can be accurately and precisely estimated using radar sensors. This illustrates the feasibility for non-contact wearable radar-based monitoring of arrhythmic risk, with particular applicability to culturally sensitive populations.

## Introduction

Sudden cardiac death (SCD) remains devastating and unpredictable worldwide [1]. The risk of SCD is higher when the QT interval adapts slowly to changes in heart rate, a phenomenon that holds also for patients with atrial fibrillation [2, 3]. These electrophysiological changes highlight how an impaired QT interval variability contributes to greater susceptibility to malignant arrhythmias and SCD [4]. The QT interval of the electrocardiogram (ECG) encompasses the complete cycle of ventricular electrical activation and repolarization, and serves as a gauge of cardiac electrical function [5]. However, the QT interval varies with heart rate [6] and with individual cardiac structural differences [7], which can obscure transient beat-to-beat repolarization irregularities [8]. The QT variability index (QTVI) quantifies beat-to-beat fluctuations in QT relative to heart rate variability (HRV), and associates with SCD risk after myocardial infarction [9–11].

The QTVI transforms the QT interval from a static measure to a dynamic one, where a high QTVI indicates disproportionately elevated repolarization variability relative to heart rate variability. QTVI has been comprehensively validated as a high-value prognostic marker in post-infarction patients in the MADIT II study, where an elevated QTVI was associated with an 80% increased risk of ventricular fibrillation [12]. Furthermore, in patients with heart failure in the GISSI-HF trial, 24-hour QTVI was a strong indicator of cardiovascular mortality (hazard ratio 4.4) [13]. Consequently, QTVI offers a direct window into the temporal instability of ventricular repolarization.

Radar sensing using either continuous-wave (CW) or frequency-modulated continuous wave (FMCW) approaches has played an emerging role in the long-term non-invasive non-contact detection of heart rate [14–18]. HRV metrics obtained from radar sensors have shown a close agreement with those from ECG [19]. In principle, radar detects movements of tissue, and relatively low frequency radar (less than 1000 MHz) can detect cardiac motion at depths of up to 20 cm under the skin. Advanced algorithms can analyse such movements to localize the exact timing of beats, constructing a reliable series of the intervals between each beat necessary for calculating measures of HRV [20].

Importantly, Australian Aboriginal and Torres Strait Islander peoples, much like many first nations peoples worldwide, experience cardiovascular conditions earlier and more severely due to structural inequities, not biological differences [21]. Indigenous peoples may be culturally sensitive to body exposure, thus making non-contact monitoring a particularly attractive technology. This study adopts a strength-based approach, recognising that emerging wearable radar technologies have potential to support Indigenous health sovereignty, and ultimately seeks to enable culturally safe, non-contact, and community-controlled cardiac monitoring.

While radar-derived HRV has been well described and studied, the translation of ECG measures of myocardial repolarization instability to non-contact methods like radar remains an unexplored area. Therefore, the first aim of the study was to compare QTVI with the analogous J-point-to-T-end variability index (JTVI) in ECG data from patients both in sinus rhythm and with atrial fibrillation. Given that the QRS duration does not vary markedly over time, we hypothesized that the JTVI and QTVI would show close agreement. The subsequent aim of the study was to bridge the gap between the timing of ECG electrical events to radar-based mechanical events. We hypothesized that it would be possible to develop an accurate and precise radar-based measure that closely agreed with JTVI by ECG.

## Methods

Ethical approval for this study was obtained from applicable local institutional human research ethics committees. In alignment with the Maiam Nayri Wingara Indigenous Data Sovereignty Principles, it is acknowledged that physiological data are not only informational but relational, representing people, communities, and environments.

### Comparing QTVI and JTVI

Single lead (lead I) ECG signals recorded for 120 seconds from two publicly available datasets (MIMIC PERform AF [22] and Erlangen hospital [23]) were evaluated. Out of 35 critically ill patients (19 with atrial fibrillation) available from MIMIC PERform AF, the first 9 with atrial fibrillation and 11 with sinus rhythm were included. Additionally, out of 30 healthy volunteers in the Erlangen hospital database, the first 20 volunteers were selected, yielding a total of 40 subjects for analysis. For each recording, beat-to-beat QT and JT intervals were extracted using automatic delineation of fiducial points (QRS start [Q], QRS peak [R], QRS end [J], and T end [T]) from the ECG signal. The standard deviation of each duration (QT, JT) was calculated. The QTVI is the normalized index of QT variability, and was computed as previously described in [10], as follows:

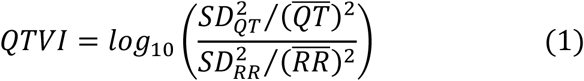

where SD_QT_ is the standard deviation of the QT interval, 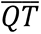 is the mean QT interval, SD_RR_ is the standard deviation of the R-R interval, and 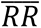 is the mean R-R interval. By analogy, the JTVI was computed as follows:

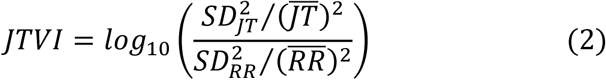

where SD_JT_ is the standard deviation of the JT interval, 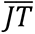 is the mean JT interval, SD_RR_ is the standard deviation of the R-R interval, and 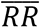 is the mean R-R interval.

### Radar data acquisition and signal processing

A continuous wave (CW) radar operating at 875 MHz was used for data collection, with the frequency chosen to create a compact, low-powered, and wearable radar system [24]. The components and size of the sensor are shown in Figure 3a. As shown in Figure 3b the experimental setup included the radar antennae on the chest (blue circles) and ECG electrodes on the wrists (red circles). Data were collected for a duration of 2 minutes in a seated position, and both radar and a single-lead ECG (lead I) were recorded simultaneously (Figure 3c and 3d). The radar signal was first detrended to eliminate DC offsets and baseline drift. A fast Fourier transform was then used to ascertain the predominant spectral components of the heart rate (*F_HR_*) and respiratory rate (*F_resp_*). The specified frequencies guided the design of subject-specific bandpass filters [F_low_, F_high_] using a modified version of the algorithm presented in [25], as follows:

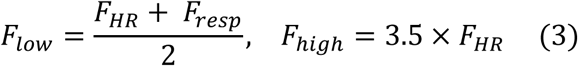

The filter was then applied to the signal to isolate the heart rate signal from the respiratory rate signal and suppress noise. A fourth-order Butterworth bandpass filter (2-45 Hz), a 50 Hz Notch filter, and a moving average filter (W=0.05×F_s_, F_s_ = sampling frequency) were also applied to the ECG signal to suppress noise, including 50 Hz electrical power noise.

Although the moving average filter introduces some time lag to the signal, it does not affect the accuracy of detecting time duration intervals as such. The raw and filtered radar and ECG signals, as well as the aligned waveforms, are shown in Figure 3e–3g, respectively. The overall signal processing block diagram is presented in Figure 3h. Figure 3i shows three beats of the radar and ECG signals, along with all the detected timepoints and durations. The diagram highlights both the obvious and non-obvious radar notches, which are discussed further below.

### Measuring the radar analogue of the JT interval

It was empirically observed that the time interval between the peak and notch of the radar signal (peak-to-notch [PN] interval) appeared to correspond to the JT interval of the ECG. Consequently, a PN detection algorithm was designed to detect these two landmarks.

The step-by-step mathematical description for identifying the peak and the notch and calculating the peak-to-notch interval of the radar signal is detailed in the Supplementary material. In summary, following filtering, the radar signal was subjected to a peak detection function. The peak detection employs the temporal constraint to ensure a physiologically meaningful detection. This constraint was defined as the minimum interval between peaks, which was calculated for each subject is based on their heart rate determined from frequency analysis (step 2 in the algorithm). After the identification of the peaks, a search window was defined for notch detection, ranging from 20-50% of the mean peak-to-peak interval following each radar peak. These search windows are indicated by the shaded regions in Figure 3i.

The algorithm for notch detection was designed to detect either an obvious or a non-obvious notch in each search window. An obvious notch is a clearly visible dip in the amplitude of the radar signal within the search window. The temporal location of this notch is determined by identifying the zero-crossing point of the first derivative of the signal. In the absence of an obvious notch, the algorithm estimates the temporal position of the non-obvious notch by detecting the peak of the second derivative of the signal. The obvious and non-obvious notches, along with their alignments with the derivative signals, are illustrated in Figure 3i.

With the radar peaks and notches identified, the PN interval was measured as the interval between each respective peak and its subsequent notch.

### Measuring the JT interval by ECG

The step-by-step mathematical description for identifying the J point and the T_end_ by ECG and calculating the JT interval is detailed in the Supplementary material. First, the R wave was detected using a peak-finding function, similar to that used for the radar signal. These points were then used as temporal references for detecting the J-point and T_end_ by identifying the zero-crossing point of the signal’s first derivative. The JT interval was calculated as the duration between each respective J point and the T_end_.

### Peak-to-notch variability index by radar

This study introduces peak-to-notch variability and the peak-to-notch variability index (PNVI) from radar as analogous measures of JT variability and the JTVI from ECG. PN variability was defined as the standard deviation of all PN intervals. The PNVI was subsequently calculated in analogy to the calculation of the QTVI and JTVI as follows:

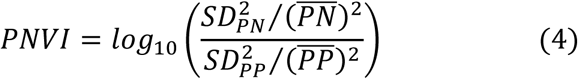

where SD_PN_ is the standard deviation of the PN interval, 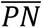 is the mean PN interval, SD_PP_ is the standard deviation of the peak-to-peak interval, and 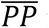 is the mean peak-to-peak interval.

### Statistical analysis

All signal processing and statistical analyses were conducted using in-house developed software in MATLAB (R2024a). The software was used for pre-processing, filtering, and feature extraction of the acquired signals, as well as for implementing statistical and validation analysis. For the radar validation component of the study, radar and ECG signals were simultaneously recorded using MATLAB R2024a environments. Correlations were described using Pearson’s correlation coefficient (r) and reported as their square (R²). Differences between methods were calculated as the mean±SD in absolute units and percentage, and described graphically using Bland-Altman analysis. A p-value<0.05 was considered statistically significant.

## Results

### Subject characteristics

The correlation cohort for determining the relationship between QTVI and JTVI by ECG consisted of patients (n=40), 23% with atrial fibrillation, heart rate 85±12 beats/min, QTVI −0.36±0.75, JTVI −0.10±0.75. No further demographic information was available for that cohort. The radar validation cohort consisted of healthy volunteers (n=20), with an age of 43±11 years, 25% were female, heart rate 66±14 beats/min, and JTVI - 0.52±0.51.

### QT and JT variability

The correlation cohort was comprised only of ECG data from two online databases and was used to validate the association between QTV and JTV. The results illustrate an excellent agreement between the standard deviations of the QT and JT intervals (R^2^=0.99, bias −0.4±1.5 ms, 1.1±3.9%), see Figure 2. These results confirm that JTV closely aligns with QTV, supporting the view that QTV is predominantly caused by fluctuations in the JT interval rather than the changes in the QRS complex.

**Figure 1:**
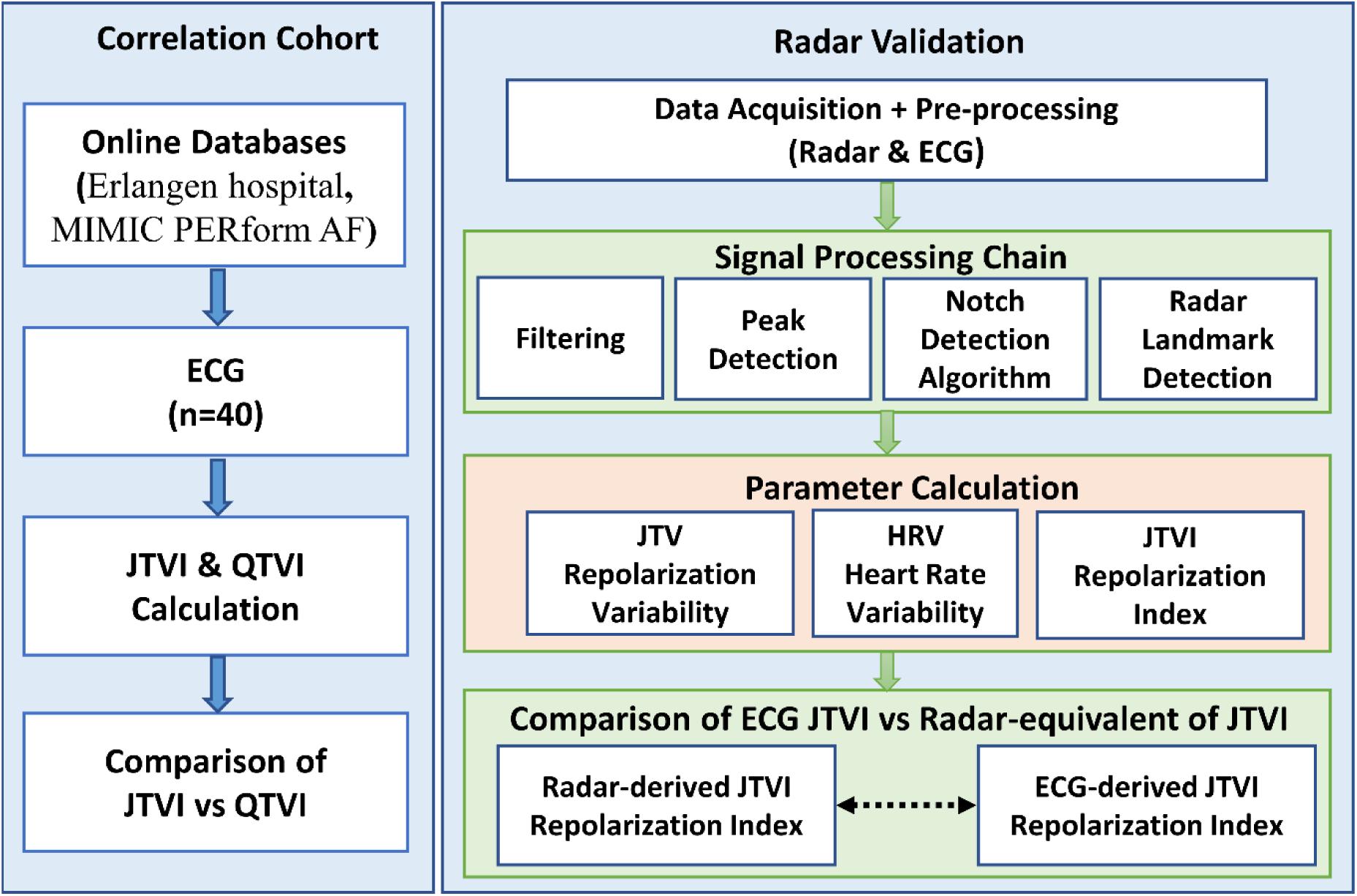
Research framework for the validation of radar system in assessing cardiac repolarization instability. The left panel illustrates the *correlation cohort* for evaluating the JT variability index (JTVI) utilizing existing online databases. The right panel illustrates the *radar validation cohort* involving filtering, peak detection, and detection algorithm to extract precise cardiac timing as well as parameter calculation and statistical analyses.

**Figure 2:**
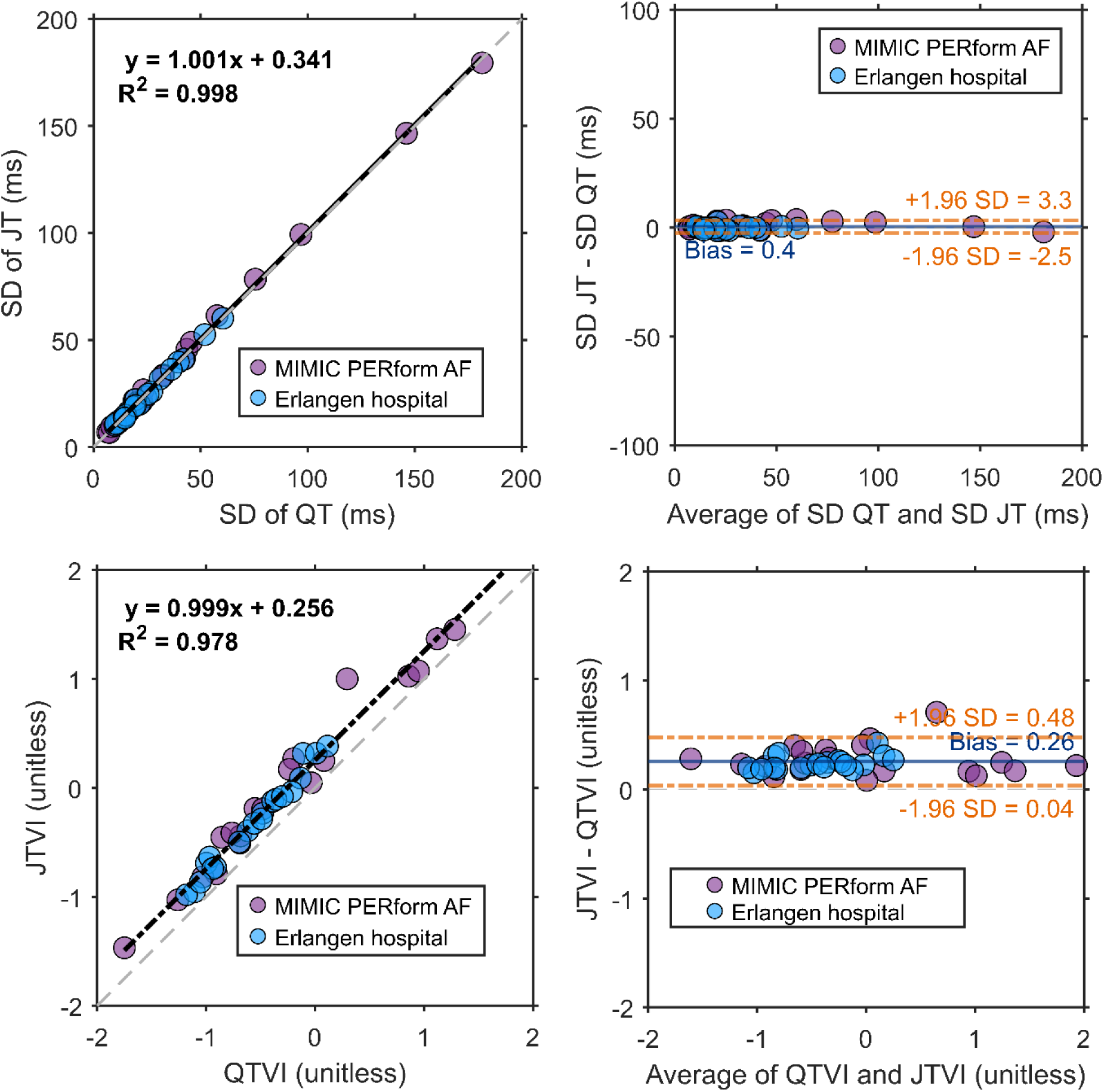
Correlation and Bland–Altman agreement analyses for pairwise QTV vs. JTV and QTVI vs. JTVI. Left panels display correlation analyses. The purple and blue dots represent data points from two datasets, the black dashed lines indicate the fitting line, and the red dashed lines display the identity lines All right panels display Bland–Altman analyses. Horizontal (blue) lines indicate the mean difference between the two parameters. Horizontal orange dotted lines indicate the upper and lower limits of agreement.

**Figure 3:**
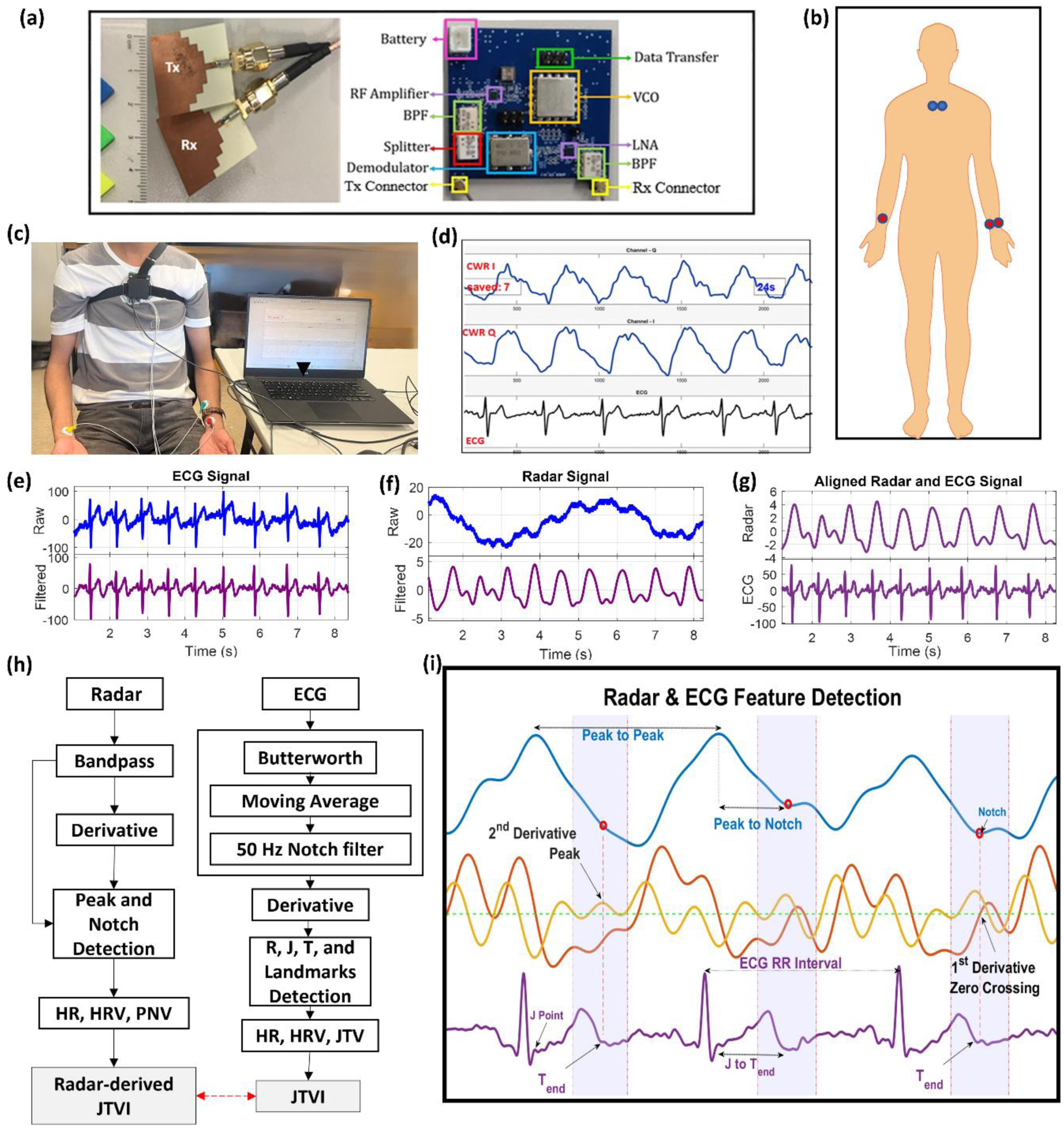
Pipeline for the radar sensing and signal processing setup. (a) Radar module components and physical dimensions, including transmitting (Tx) and receiving (Rx) antennae, radio frequency (RF) amplifier and bandpass filter (BPF), demodulator, voltage control oscillator (VCO), low noise amplifier (LNA). (b) Placement of non-contact radar antennae (blue dots) and ECG skin electrodes (red dots) on the body. (c) Experimental data collection setup. (d) Signal recording interface display illustrating the continuous wave radar (CWR) in- phase (I) channel and quadrature (Q) channel and ECG. (e) Example of acquired raw and filtered ECG signals. (f) Example of raw and filtered radar I/Q signal. The radar I/Q signal is selected from either the I or Q channel that demonstrates the optimal morphological pattern. (g) Temporally aligned display of simultaneously acquired and filtered radar and ECG waveforms. (h) Overview of the signal processing and landmark detection algorithms including heart rate (HR), heart rate variability (HRV), radar peak-to-notch variability (PNV), PNV index (PNVI), J-to-T-end variability (JTV), and JTV index (JTVI). (i) Radar and ECG feature detection, including the radar signal, its first and second derivatives with respect to time, and the simultaneous ECG signal, showing annotated points of interest (peak, notch, J, and T_end_) and corresponding intervals (Peak-to-Peak, R–R, Peak-to-notch). Red circles denote detected notches in the radar signal. Obvious notches, visible as clear dips, are detected via first-derivative zero-crossings within a search window. If an obvious notch is absent, the notch is determined as the peak of the second derivative to capture subtle slope changes. The purple shaded area denotes the range of the search window for the notch, defined as 20-50 % of the peak-to-peak interval.

### QTVI and JTVI

Furthermore, the JTVI exhibits a strong correlation with QTVI. The correlation plot reveals a strong linear correlation (R²=0.98, QTVI=1.0*JTVI−0.26) and a small and consistent bias (0.26±0.15), which would be expected given the systematic differences between JT and QT, where the JT interval accounts for approximately 70–80% of the QT duration [3]. The median JT/QT ratio of 0.75 suggests a theoretical difference of −0.25 between JTVI and QTVI, which closely aligns with the observed mean bias (−0.26). This close agreement suggests that QTVI can be reliably approximated from JTVI, simplifying the analysis of ventricular repolarization variability without significant loss of accuracy.

### Radar analogues of JT and JTVI

The radar-based system successfully delineated the peak-to-notch interval as an analogue to JT interval of ECG. Figure 4 demonstrates the temporal relationship between the JT interval and peak-notch interval for two participants. Both participants demonstrate consistent heartbeat timings, with the JT and peak-notch intervals maintaining stable alignment in their respective cardiac cycles. These findings reveal a stable, predictable link between the heart’s electrical signals and its mechanical movements, confirming that the detection method is reliable. Quantitatively, radar accurately estimated JTV (R²=0.88, bias 0.1±4.9 ms, 6.4±2.7 %), HRV (R²=0.93, bias 16.0±22.1 ms, 2.4±2.8 %), and JTVI (R²=0.99, bias 0.00±0.05, 0.0±6.5 %), see Figure 5. The excellent agreement between ECG and radar-estimated JTVI (PNVI) illustrates that the combined metric, which integrates both repolarization and heart rate dynamics, can be accurately computed from non-contact radar data. Across all parameters, the results show a strong alignment between radar-derived measurements and ECG reference values. These results validate the ability to quantify myocardial repolarization instability using a wearable radar system.

**Figure 4:**
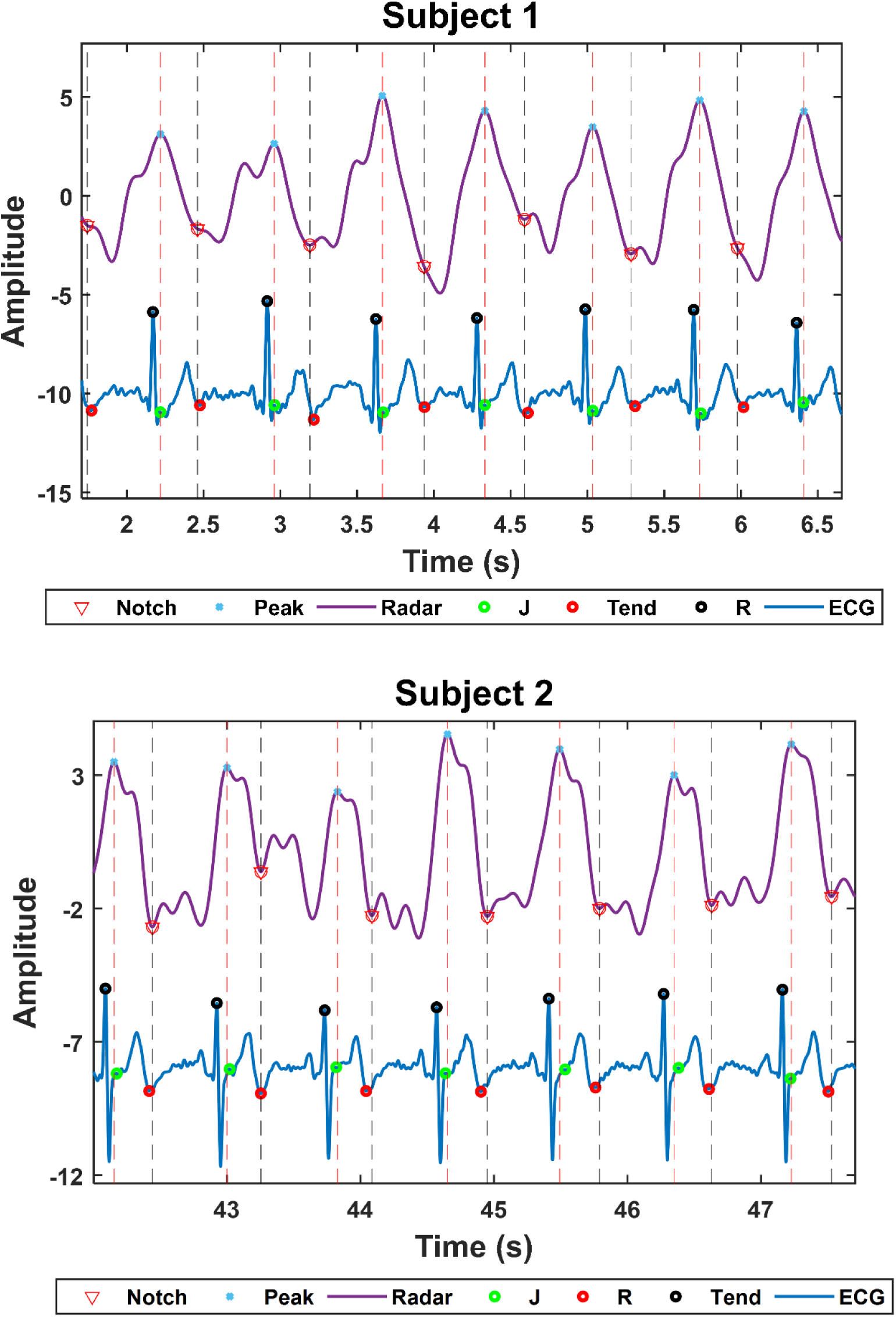
Examples of radar (purple trace) and ECG (blue trace) data for two representative subjects, where red and grey dashed vertical lines denote the automatically detected radar peak (blue solid circle) and notch (red open circle), respectively. The time interval between two consecutive red and grey lines denotes radar-derived peak-to-notch interval, which shows good visual agreement with the interval between the automatically detected J point (green open circle) and the T_end_ (red open circle) of the ECG.

**Figure 5:**
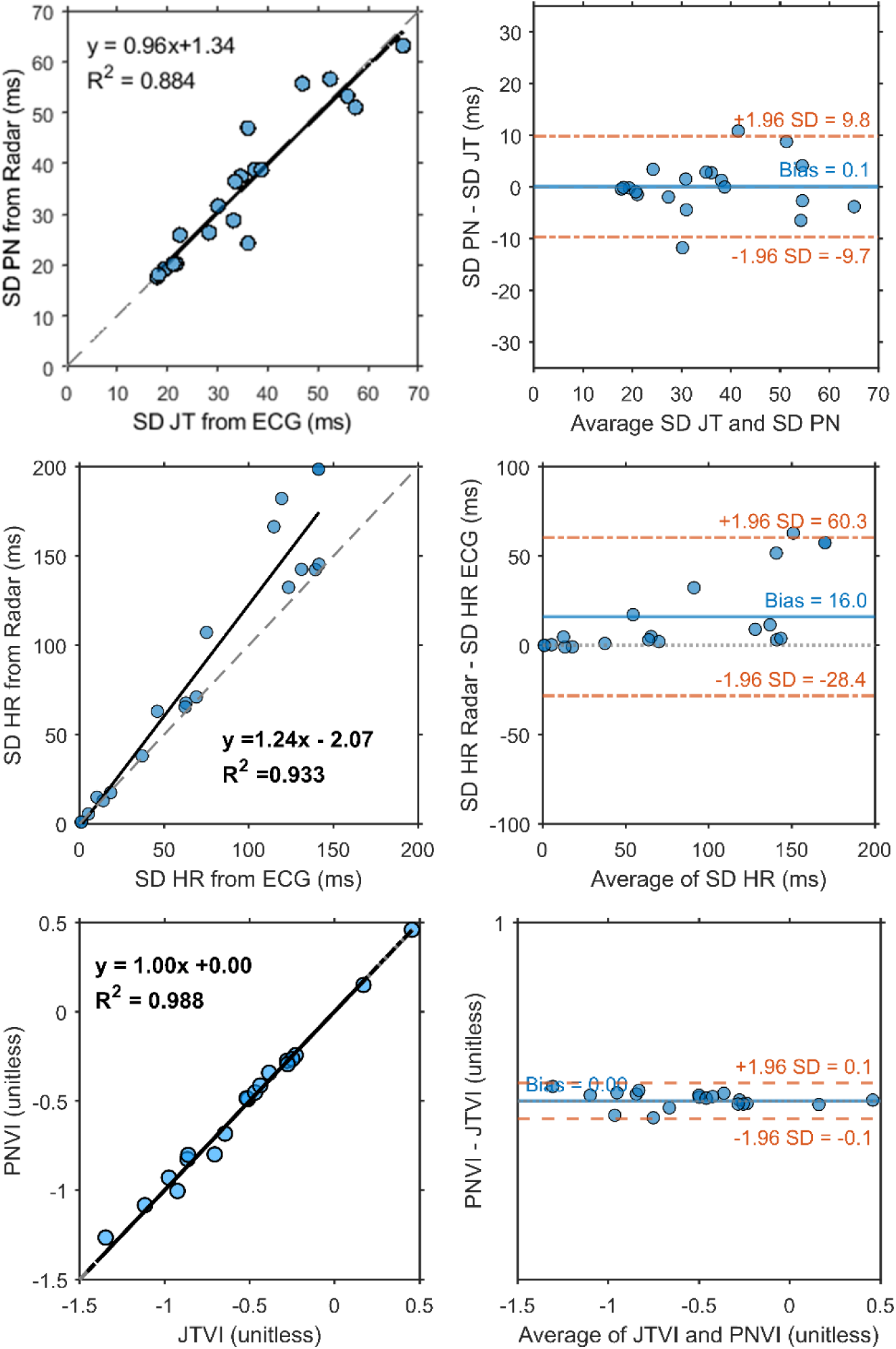
Correlation and Bland–Altman plots comparing radar-derived cardiac metrics (standard deviation of the peak-to-notch [PN] intervals, standard deviation of normal beat-to-beat intervals [SDNN], and peak-to-notch variability index [PNVI]) against reference ECGmeasurements (standard deviation of JT, SDNN and J-to-Tend variability index [JTVI]). The grey dotted lines indicate the identity line, and the red dashed lines display the line of linear correlation. Horizontal (blue) lines indicate the mean difference between the two measures. Horizontal orange dotted lines indicate the upper and lower 95% limits of agreement

## Discussion

The main findings of our study were that QTVI and JTVI adjusted for a small bias are effectively interchangeable for quantifying myocardial repolarization instability, and that JTVI can be accurately and precisely estimated using radar sensors.

### JTVI and QTVI

The QRS complex remains stable across cycles, confirming that QT interval variability is largely driven by changes in the JT segment. Analysis of beat-to-beat intervals from 40 patients revealed a strong correlation between SDs of JT and QT intervals. The data showed a near-perfect linear correlation with excellent precision. Furthermore, the normalized indices JTVI and QTVI showed remarkable similarity, separated by only a small and systematic difference. This small bias aligns with theory, given that the JT interval makes up about three-quarters of the total QT length. In practice, measuring the JTVI provides a highly accurate and simple way to assess repolarization instability, capturing the same core information as QTVI. Previous literature has examined the use of JT and JTVI obtained from ECG signals. These repolarization-specific metrics correlate strongly with QT-based markers such as QTVI. For example, children with a ventricular septal defect exhibited prolonged heart-rate corrected QT while JT remains unchanged, highlighting the superiority of JT for assessing true repolarization [26]. In addition, the JT_peak_ variability index (JT_p_VI), which represents the early phase of repolarization, has been shown to have a strong correlation with QTVI (R^2^=0.73) and an even stronger correlation with JTVI (R^2^=0.80) [27].

### Estimating JTVI with radar

In addition, the work focused on developing a non-contact method for JT interval estimation. A compact, wearable radar sensor was developed to enable mechanical detection of ventricular repolarization timing. The principal idea is that the radar signal reflects the heart’s mechanical activity. The main radar peak is thought to correspond to the maximum volume and is empirically aligned with the J-point of the ECG, while a subsequent notch is thought to corresponds to the end of systole and is empirically aligned with the conclusion of T wave (T_end_) by ECG. Taken together, the detection of these two radar landmarks and the interval between them provides a temporal estimation of the repolarization phase of the ECG. A developed processing pipeline was implemented to extract this interval reliably, beginning with the identification of distinct radar peaks.

Following this, the peak-to-notch detection algorithm identified the notch as either a signal dip or an inflection point, yielding reliable radar-based repolarization timings validated against ECG. The radar-derived PNVI demonstrated an excellent agreement with ECG-derived JTVI.

Radar-based sensing has evolved substantially, with successful translation of cardiorespiratory parameters into non-contact measurements [18, 28]. The radar-based successful detection of respiratory patterns [29], heart rate [30], and HRV [31] has previously been demonstrated, showing a strong agreement between radar-derived metrics and the ECG. The advancements in radar sensing have further refined this capability. For instance, 77-81 GHz FMCW radar has been shown to achieve exceptional R-R interval accuracy (bias 0.02 ms) [32], while a mmHRV system has been used for continuous HRV monitoring [20].

Beyond laboratory validation, radar has been shown to be capable of operating effectively in clinical settings [33].

Prior studies have largely focused on vital signs and generic signal morphology without linking mechanical signatures to electrophysiological markers. Further research has started probing deeper into cardiac mechanics and identifying radar-derived pulse waveforms to capture distinct aspects of cardiac function [34]. However, a critical gap has been the inability to measure cardiac ventricular repolarization through a non-contact approach. Ventricular repolarization measures such as QT interval, QTV, and QTVI have been considered important for quantifying ventricular repolarization instability and providing a valuable estimate of SCD risk [2, 3, 13]. The results of the current study bridge this gap by identifying radar-based analogues corresponding to the J-point and T_end_ of the ECG. This is the first non-contact estimation of JTVI through wearable radar. Moreover, we demonstrate that JTVI is strongly correlated to QTVI. This represents an important advancement in non-contact sensing beyond vital sign monitoring and into the domain of electrophysiological risk assessment.

### Indigenous health considerations

Beyond technical performance, this work demonstrates wearable radar sensing that can be integrated within frameworks of Indigenous-led health innovation. This is particularly valuable given that some Indigenous peoples may be culturally sensitive to body exposure, thus making non-contact monitoring a particularly attractive technology. When embedded within Aboriginal governance systems, non-contact monitoring offers a pathway for self-determined care, where communities manage data, interpretation, and response on their own terms. The concept of Kaadaninny (deep listening) offers a guiding ethic for such translation, wherein innovation should move at the pace of trust and in the rhythm of relationship. These principles ensure that technological advancement strengthens, rather than supplants, cultural systems of health knowledge. From an Indigenous governance perspective, the translation of cardiovascular innovation into clinical or community contexts must extend beyond accuracy and efficiency to include sovereignty and benefit. Aboriginal and Torres Strait Islander communities have long engaged in forms of embodied listening and monitoring the rhythms of body and Country through deep observation and care. A strength-based lens positions technologies like wearable radar cardiography as companions to these knowledge systems, not replacements. By aligning design with Indigenous Data Sovereignty and self-determined health priorities, such innovations can enhance equitable access to early cardiac risk assessment across remote and urban communities, and provide a transferable model for Indigenous and under-served populations globally.

### Limitations

Despite promising results, the current study still has limitations. Primarily, the validation was carried out with a relatively small, homogenous cohort of healthy individuals, and this limits the generalizability of the system. Moreover, short recording were obtained under controlled and seated conditions, and further validation is required to determine the performance of this approach under ambulatory and dynamic conditions. The current signal processing pipeline was optimized for normal conditions, and modifications will likely be required to either ensure motion artifacts suppression for data processing during normal activities, or to limit analysis to data acquired at rest, for example by detecting rest using an integrated accelerometer. Future work should evaluate system performance in broader, and more diverse clinically relevant populations, including patients with cardiac conditions such as heart failure or myocardial infarction that exhibit repolarization instability. Future investigations should also be performed during normal daily activities to monitor cardiac repolarization under ambulatory and dynamic conditions. Advanced adaptive filtering techniques will be crucial for enhancing the resilience of the signal processing algorithm and suppressing motion artifacts associated with daily-life monitoring scenarios. Another direction for future work will be estimating the prognostic value of radar-derived intervals for predicting cardiac arrhythmias.

## Conclusion

This study introduces a new approach for monitoring ventricular repolarization dynamics using a wearable radar-based system. It proposes and validates an estimation of JTVI derived from mechanical cardiac activity measured by a radar sensor. The work demonstrates an excellent correlation between JTVI and QTVI in ECG signals, establishing JTVI as a surrogate metric for QTVI after correcting for a small bias. Subsequently, the radar-derived PNVI demonstrates excellent agreement with the JTVI by ECG. To the best of our knowledge, this is the first radar-based foundational framework for quantifying myocardial repolarization instability. This investigation paves the way for radar-based evaluation of cardiac arrhythmic risk, a promising direction for non-contact health monitoring technologies that may be particularly applicable to culturally sensitive populations.

## Data Availability

All data produced in the present study are available upon reasonable request to the authors

## Acknowledgements

This research was supported by an Australian National Health and Medical Research Council (NHMRC) Development Grant (2040091), with an expressed aim of developing novel non-contact methods to monitor and improve health for first nations peoples. The research was conducted on the unceded lands of the Wurundjeri Woi Wurrung people of the Kulin Nation, in Naarm (Melbourne), Australia. The authors acknowledge Elders past and present and recognise the Whadjuk Noongar and Gadigal (Eora Nation) Countries as home to members of the research team. The authors honour the enduring custodianship of knowledge, health, and observation that these lands embody. In the spirit of global reciprocity, the authors also acknowledge the Sámi peoples of northern Europe, whose sustained relationships to land, rhythm, and environment remind us that attunement to the body’s signals is part of a wider human practice of listening. TE is a Whadjuk Noongar researcher whose work focuses on Indigenous research governance, health sovereignty, and the ethical translation of science into community benefit. His contribution to this project centres on integrating relational accountability and Indigenous Data Sovereignty within biomedical innovation. Future applications of wearable radar sensing in Aboriginal and Torres Strait Islander contexts will be governed by Indigenous data governance frameworks to ensure that ownership, access, and interpretation remain community controlled. The authors’ collaborative approach reflects the principle of Kaadaninny (deep listening) where research integrity is measured not only by technical accuracy but by the quality of relationships among researchers, participants, and the knowledge systems they engage. The authors acknowledge a model of relational science in which cultural ethics and technological precision are interdependent.

## Supplementary material

### Algorithm 1: Peak to Notch (PN) Duration Calculation

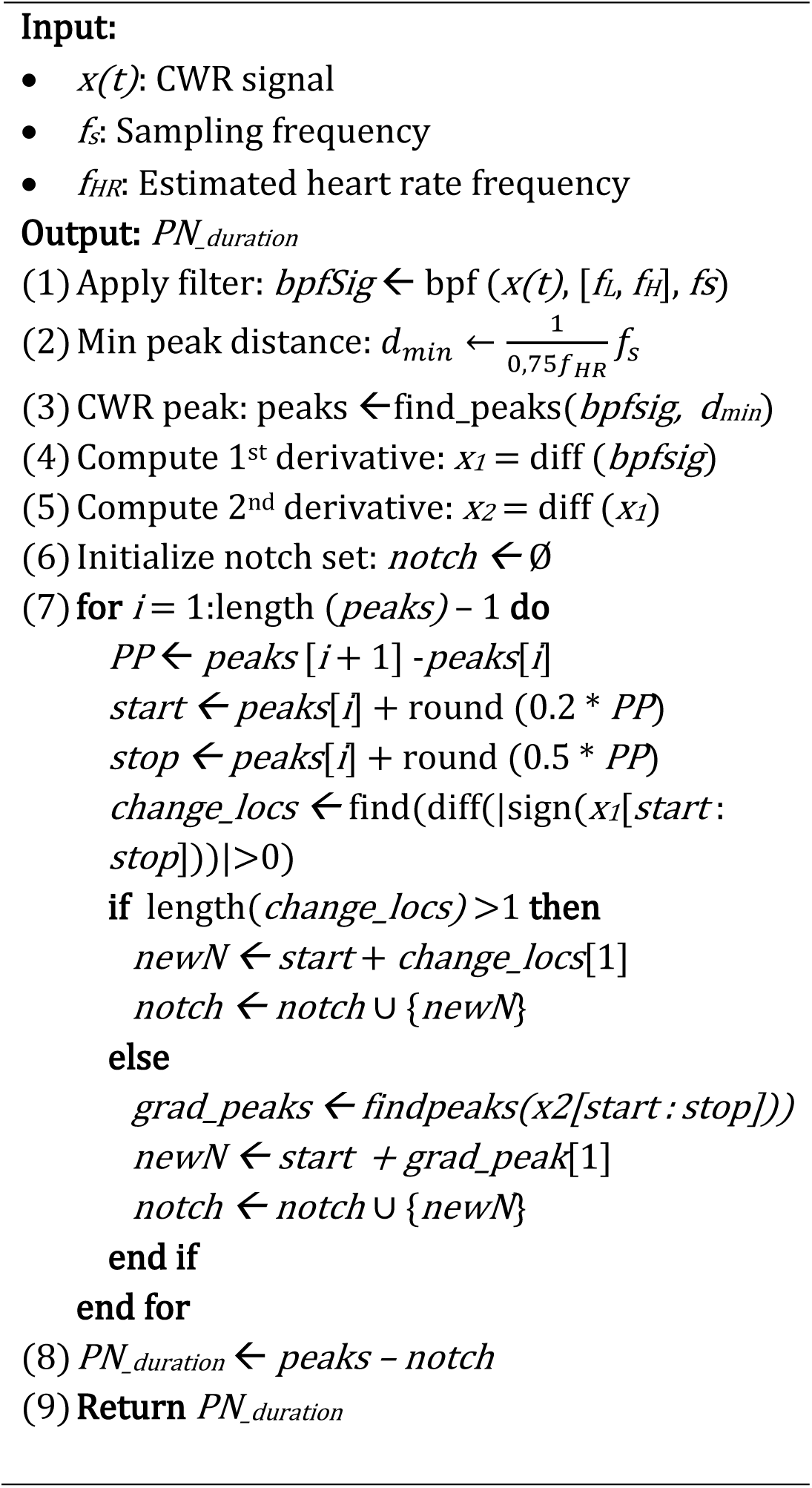

### Algorithm 2: JT Duration Calculation

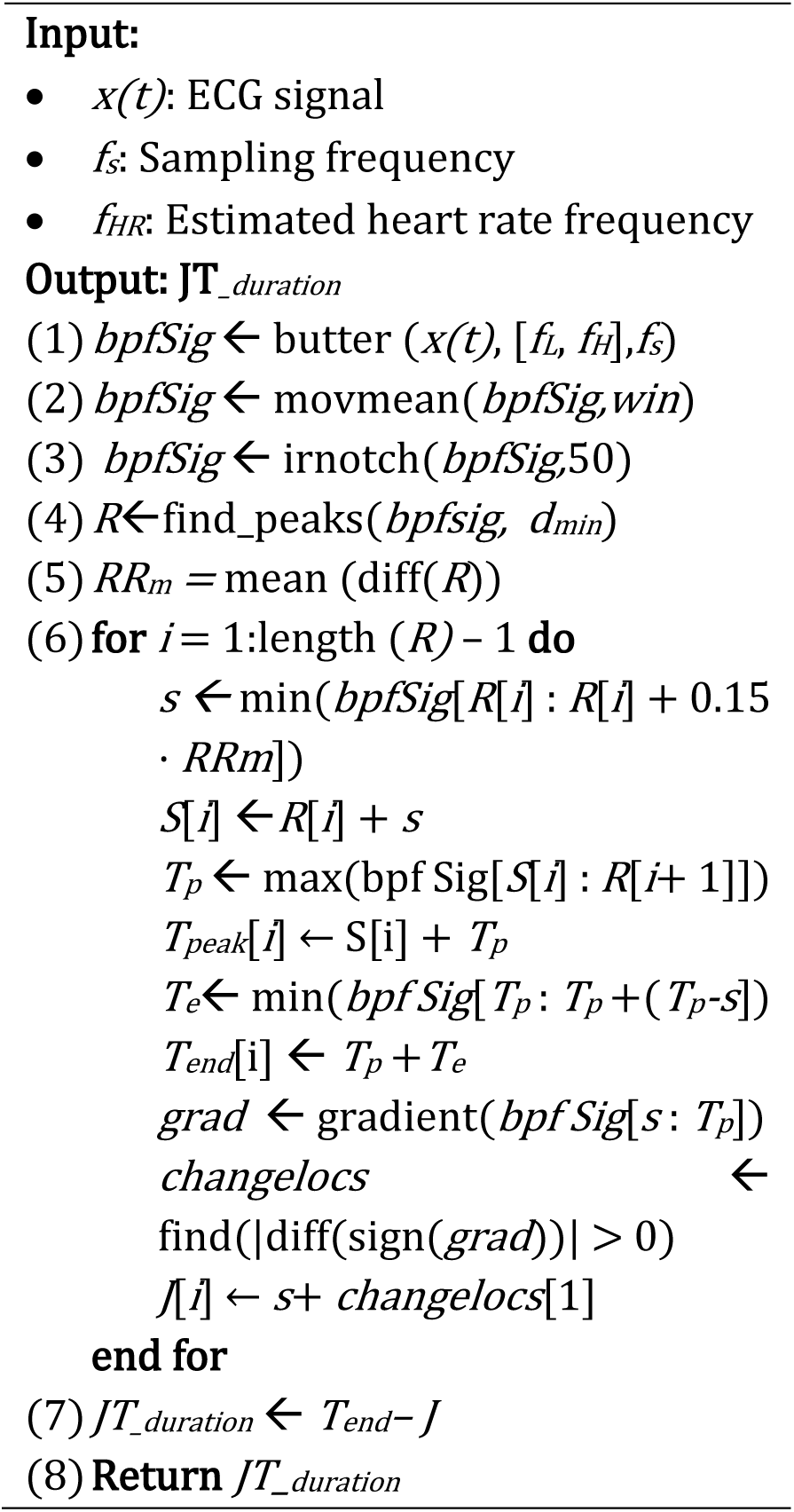

